# Detection of SARS-CoV-2 in saliva using tailed amplicon sequencing

**DOI:** 10.1101/2021.05.19.21257217

**Authors:** Aaron Garoutte, Tasha M. Santiago-Rodriguez, Heather L. Fehling, Rafal Iwasiow

**Affiliations:** Diversigen, Inc., Houston, TX, USA, 77046; Clinical Reference Laboratory, Inc. Lenexa, KS 66215; DNA Genotek, Inc., Ottawa, Ontario, CA, K2V 1C2

**Keywords:** genomic surveillance, SARS-CoV-2, sequence capture, tailed amplicon sequencing

## Abstract

The most recent virus from the *Coronaviridae* family infecting humans, SARS-CoV-2, has resulted in a global pandemic. As part of the surveillance efforts, SARS-CoV-2 genomes are increasingly being made publicly available. Methods that include both short- and long-read sequencing have been used to elucidate SARS-CoV-2 genomes; however, many of these untargeted approaches may require deeper sequencing for greater genome coverage. For this reason, sequence capture or amplicon-based approaches for SARS-CoV-2 genome sequencing have been developed. The present study evaluated a modified sequence capture approach, namely, tailed amplicon sequencing, to determine SARS-CoV-2 near complete genome sequences from the saliva of infected individuals. Particularly, the suitability of saliva samples stored at room temperature using OMNIgene^®^•ORAL OME-505 was evaluated. The tailed amplicon sequencing approach poses the additional advantage of being a cost-effective method for library preparation. Different known SARS-CoV-2 variants were identified across the infected subjects, with an average of > 99.4% genome coverage. This methodology also enabled robust genomic surveillance using phylogenetic analyses. The present study supports the suitability of saliva stored at room temperature using collection devices for SARS-CoV-2 variant detection. Importantly, the present study supports the use of tailed amplicon sequencing approaches as an alternative, cost-effective method for SARS-CoV-2 detection in saliva for genomic surveillance.

## INTRODUCTION

Human saliva is home to over 700 microbial species, and serves as protection against bacterial, fungal and viral infections that could potentially reach the respiratory tract (Kilian et al., 2016). It is composed of water and the secretion of salivary glands, dental plaque, as well as nasal and bronchial secretions; thus, several different viruses in human saliva can be identified, some of which may cause disease. The interactions with viruses and specific saliva components can be virus-specific, complex and can potentially influence their detection as a result of the biological functions of saliva (Y. Li et al., 2020). Viruses including, but not limited to hepatitis A (Leon et al., 2015), hepatitis B (Khadse et al., 2016; Parizad et al., 2016), cytomegalovirus (De Carvalho Cardoso et al., 2015), Epstein Barr virus (Kwok et al., 2015), Zika virus (Bonaldo et al., 2016), and Severe Acute Respiratory Syndrome-Associated coronavirus (SARS-CoV) (Wang et al., 2004), have all been detected in the saliva of infected individuals. This has prompted the investigation of saliva as a potential sample type for the diagnosis of several of the mentioned viruses, and of potential emerging and re-emerging viral pathogens.

The most recent virus from the *Coronaviridae* family known to infect humans, SARS-CoV-2 has also been identified in the saliva of infected individuals (Y. Li et al., 2020). While the origin of SARS-CoV-2 is a subject of ongoing research and speculation, it shares 96% identity with a bat coronavirus (Zhou et al., 2020), 90% with coronaviruses present in pangolins (Zhang et al., 2020), and 80% with SARS-CoV (Zhou et al., 2020). SARS-CoV-2 is known to also be transmitted through aerosol droplets, which may also include salivary droplets. Several studies have identified SARS-CoV-2 in saliva (Han and Ivanovski, 2020; Y. Li et al., 2020), and have proposed its use as an alternative to nasopharyngeal (NP) and oropharyngeal (OP) sample collection, which pose additional discomfort and the need for trained personnel (Yoon et al., 2020). In addition, with the availability of at-home collection kits, subjects are more willing to self-collect saliva(Valentine-Graves et al., 2020). Supervised, self-collected saliva has shown to perform similarly to clinician-collected NP swabs for the detection of SARS-CoV-2 in terms of virus detection and quantification (Noah et al., 2020). Success of detection methods for SARS-CoV-2 depends, among many things, on the persistence and inactivation of the virus and nucleic acids. For example, SARS-CoV-2 is detectable using reverse transcription PCR (RT-PCR) for an average of 18 to 20 days, and up 21 to 26 days in some instances (To et al., 2020; Yoon et al., 2020). Saliva has also been suitable for antibody testing, showing that SARS-CoV-2 antibodies can be detected as early as 10 days (To et al., 2020). This further supports the use of saliva for the detection of SARS-CoV-2 nucleic acids and antibodies. Other factors affecting SARS-CoV-2 detection include efficient viral lysis, with various commercially available kits and methods showing varying levels of efficiency (Chu et al., 2020).

The gold standard for the detection of SARS-CoV-2 in any sample type is RT-PCR or quantitative RT-PCR (RT-qPCR) (Takeuchi et al., 2020). As with any PCR-based method, success will depend on primer specificity; thus, the RNA target genome(s) sequence(s) should be known in order to increase sensitivity (D. Li et al., 2020). Other, less evaluated methods for the detection and surveillance of SARS-CoV-2 in various sample types include high-throughput sequencing. RNA high-throughput sequencing was originally used in combination with other methods to identify the novel coronavirus in subjects suffering from pneumonia of unknown origin, showing a similarity close to 90% to coronaviruses present in bats(Wu et al., 2020). High-throughput sequencing can continue to be used for the surveillance of known and novel SARS-CoV-2 variants, and also to determine genetic diversity, which is usually not provided by RT-PCR or RT-qPCR (Pérez Cataluña et al., 2021). Two approaches have been developed for the detection of SARS-CoV-2 in various sample types using high-throughput sequencing, namely, untargeted and amplicon-based. Untargeted high-throughput sequencing provides the advantage of detecting and monitoring both known and emerging SARS-CoV-2 variants, with the caveat of requiring deeper sequencing to obtain the needed genome coverage for identification, making this approach relatively more cost-prohibiting. Amplicon high-throughput sequencing, or sequence capture methods, have been more widely applied for the detection of SARS-CoV-2 as it represents a more cost-effective approach. One caveat of sequence capture approaches is the failure to cover the entire viral genome as the primers usually cannot cover the genomic ends (Gohl et al., 2020).

Approaches, such as the ARTIC network, have developed methods for amplicon pool preparation for the sequencing of SARS-CoV-2 which involve a ‘lab-in-a-suitcase’ that can be used in remote and resource-limited areas (https://artic.network/1-about.html). In the ARTIC network protocol, the first cDNA strand is enriched by amplifying with two different pools of primers. This generates amplicons tiling the virus genome, which are then subjected to either Illumina or Oxford Nanopore library preparation and sequencing. More recently, a modification to the ARTIC network protocol, known as tailed amplicon sequencing, has been developed to reduce library preparation cost and time (Gohl et al., 2020). The method has been shown to achieve results comparable to the ARTIC network protocol (Gohl et al., 2020). Briefly, in the tailed amplicon approach, the first cDNA strand is enriched using the ARTIC v3 primers. In this case, the primers also contain adapter tails that allow sequencing libraries to be created through a second indexing PCR. This, in turn, adds sample-specific barcodes and flow cell adapters. Notably, this modification to the ARTIC protocol has not been extensively tested in saliva samples. Therefore, the main aim of the present study was to evaluate the suitability of the modified tailed amplicon sequencing protocol for the detection of SARS-CoV-2 and its ability to discern potential variants in human saliva.

## MATERIALS AND METHODS

### Sample collection and RNA extraction

Saliva samples were collected from various geographical locations in United States using the OMNIgene^®^•ORAL OME-505 device (DNA Genotek, Inc) and shipped to Clinical Reference Laboratory, Inc. for testing using the CRL Rapid Response^™^ COVID-19 Test. After testing, the remaining saliva samples collected and stabilized in the OMNIgene^®^•ORAL OME-505 devices, were stored at room temperature for up to 15 days prior to RNA extraction (**Table 1**). At a later point, one to fifteen days post diagnosis, eight randomly selected samples from a pool of samples which had previously tested positive for SARS-CoV-2 by the CRL Rapid Response^™^ COVID-19 Test were selected to be sequenced. The only criteria used for sample selection was an initial Ct value <30 (**Table 1**). The randomly selected samples were extracted using the Zymo Quick RNA/DNA Viral MagBead kit (Cat. No. R2141) following manufacturer’s instructions.

**Table 1.**
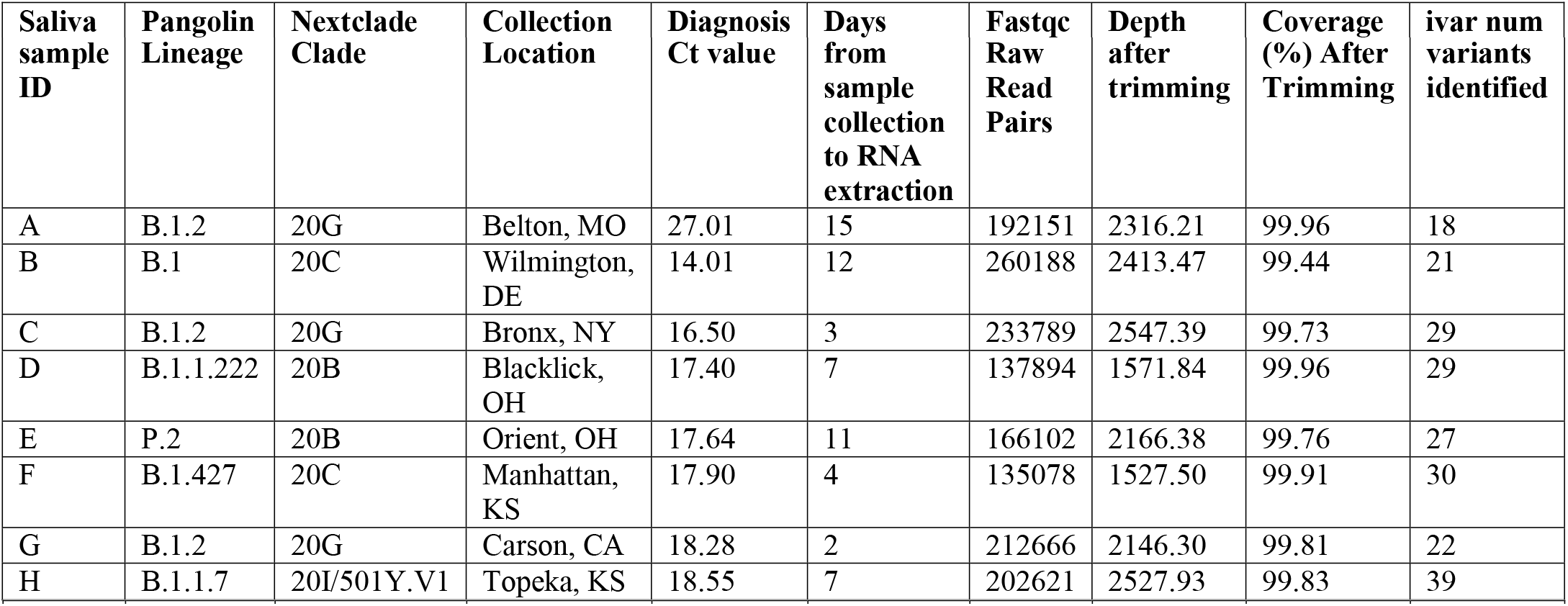
Summary of relevant metadata associated with saliva samples including: Pangolin lineage which indicates the strain identity, Nexclade clade identification which shows the phylogenetic lineage of the variant, qPCR Ct value for diagnosis, number of raw fastq reads, read depth on the genome after trimming primers, percent coverage of the genome after trimming and the number of variants (mutations) identified using iVar.

### Sample processing, sequencing and bioinformatic analyses

The extracted RNA was then prepared and sequenced using previously described methods in Gohl et al., 2020 (Gohl et al., 2020). Briefly, the integrity of the extracted RNA was analyzed as described previously. After RNA quality and integrity was checked, RT-qPCR was also performed as described previously. RNA was also processed through the amplicon-based sequencing method that utilizes adapter tails with the ARTIC network v3 primers, allowing for a more efficient library preparation (Itokawa et al., 2020). Sequencing was also performed as previously described using a MiSeq 600□cycle v3 kit following manufacturer’s instructions. After sequencing, the paired ends were joined using PANDAseq (Masella et al., 2012). Unaligned reads were aligned to Wuhan-Hu-1 SARS-CoV-2 genome (MN908947.3) using BWA (Li and Durbin, 2010; Wu et al., 2020). The Ivar software package was used for trimming and filtering reads (Grubaugh et al., 2019). Ivar was also used to call variants and generate consensus sequences. The consensus sequences were then strain typed using Pangolin (github.com/cov-lineages/pangolin). Genome coverage plots were created using custom R scripts. Phylogenetic tree was created using the Nextstrain web interface (Hadfield et al., 2018)(v0.14.2, commit: f62d906, build 655).

## RESULTS AND DISCUSSION

The present study evaluated a modified sequence capture approach, namely tailed amplicon sequencing, for the detection of SARS-CoV-2 in the saliva of infected individuals [21]. Detection of SARS-CoV-2 in the saliva of infected individuals, both symptomatic and asymptomatic, has expanded the toolbox of methods for the detection and diagnostics of the virus. While RT-PCR and RT-qPCR are the standard methods for the detection of SARS-CoV-2 in various sample types, tailed amplicon sequencing approaches are also capable of identifying SARS-CoV-2 in the saliva of infected individuals. Notably, results from the present study showed that the tailed amplicon sequencing approach was successful in the identification of various SARS-CoV-2 variants in human saliva (**Table 1**). Results also showed the suitability of OMNIgene^®^•ORAL OME-505 for sufficient SARS-CoV-2 RNA recovery for tailed amplicon sequencing. Moreover, storage of saliva samples at room temperature using OMNIgene^®^•ORAL OME-505 for up to 15 days further supports the capacity of this collection device to capture saliva composition at the time of collection, essential for SARS-CoV-2 diagnostics and surveillance. While upper and lower respiratory tract specimens were first to be recommended for SARS-CoV-2 diagnosis, saliva has gained acceptance as a suitable, non-invasive sample type. Indeed, saliva has been included in numerous FDA Emergency Use Authorizations for the purpose of SARS-CoV-2 diagnosis, including CRL Rapid Response^™^. The OMNIgene^®^•ORAL OME-505 saliva collection device, which has received EUA for SARS-CoV-2 sample collection, allows for self-collection and to circumvent the need to store samples on ice or frozen.

Interestingly, the tailed amplicon sequencing approach tested in the present study provided near complete genomes (>99.4 % average genome coverage) for all eight samples tested (**Table 1** and **Figure 1**). In addition, the tailed amplicon method produced a relatively uneven genome coverage balance. Indeed, this has been noted previously when comparing the tailed amplicon method with the ARTIC network protocol (Gohl et al., 2020). One feasible explanation for this unevenness in genome coverage is the better balance of the untailed primers utilized in the ARTIC network protocol. Nevertheless, near complete SARS-CoV-2 genomes were obtained in the present study, which may improve insights into virus mutations, evolution, and adaptation (e.g., increased transmissibility and infectivity) compared to evolutionary relationships across spike protein sequences alone. Unlike specific genes, complete or near complete genomic sequences provide the most high-resolution information that allows determination of variant and strain relatedness during outbreaks and pandemics. Thus, the development of cost-effective and less time-consuming protocols to determine genomic sequences is of importance. In addition, near complete genome information may also aid in the classification of SARS-CoV-2 variants into strains. SARS-CoV-2 strain level resolution using tailed amplicon sequencing approaches could potentially aid to bridge mutations within SARS-CoV-2 genomes at a global scale, which in turn may aid to understand virus transmission and population dynamics. Variant level resolution of SARS-CoV-2 may also facilitate the identification of novel target regions for vaccine development and therapeutics, particularly regions across SARS-CoV-2 genomes that may be shared across variants and potential strains.

**Figure 1.**
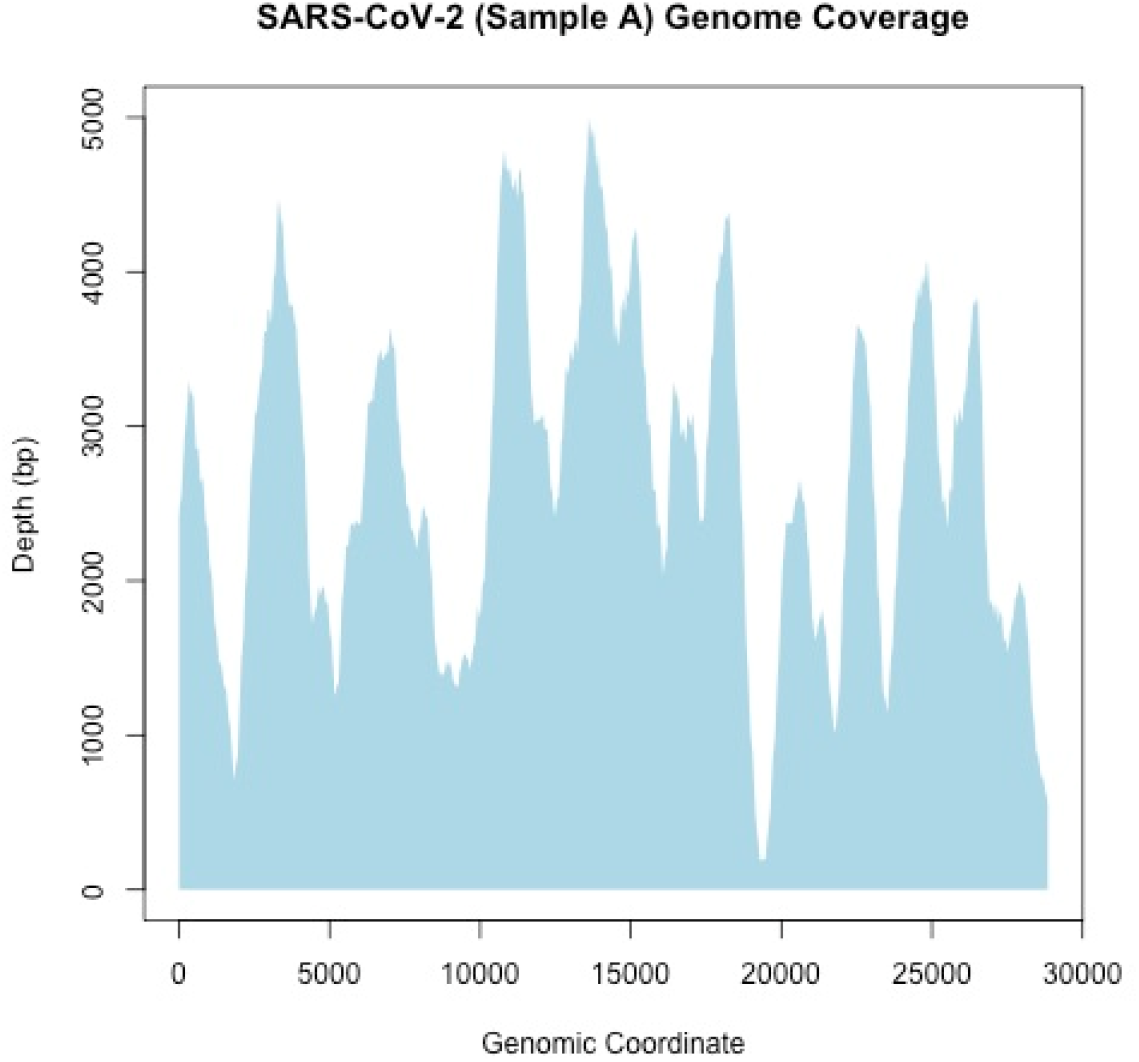
Representative example of SARS-CoV-2 genome coverage. Graph illustrates that the combination of sample collection and sequencing methodology is sensitive enough to capture most of the genome with very high coverage.

Near complete genomic sequences obtained using the tailed amplicon sequencing enabled deciphering phylogenetic relationships across the various SARS-CoV-2 variants identified in the saliva of the infected individuals in comparison with known clades (**Figure 2**). Results show that, overall, the tailed amplicon method described above is sensitive enough to resolve, not only large-scale genomic differences across different clades, but also differences within clades.

**Figure 2.**
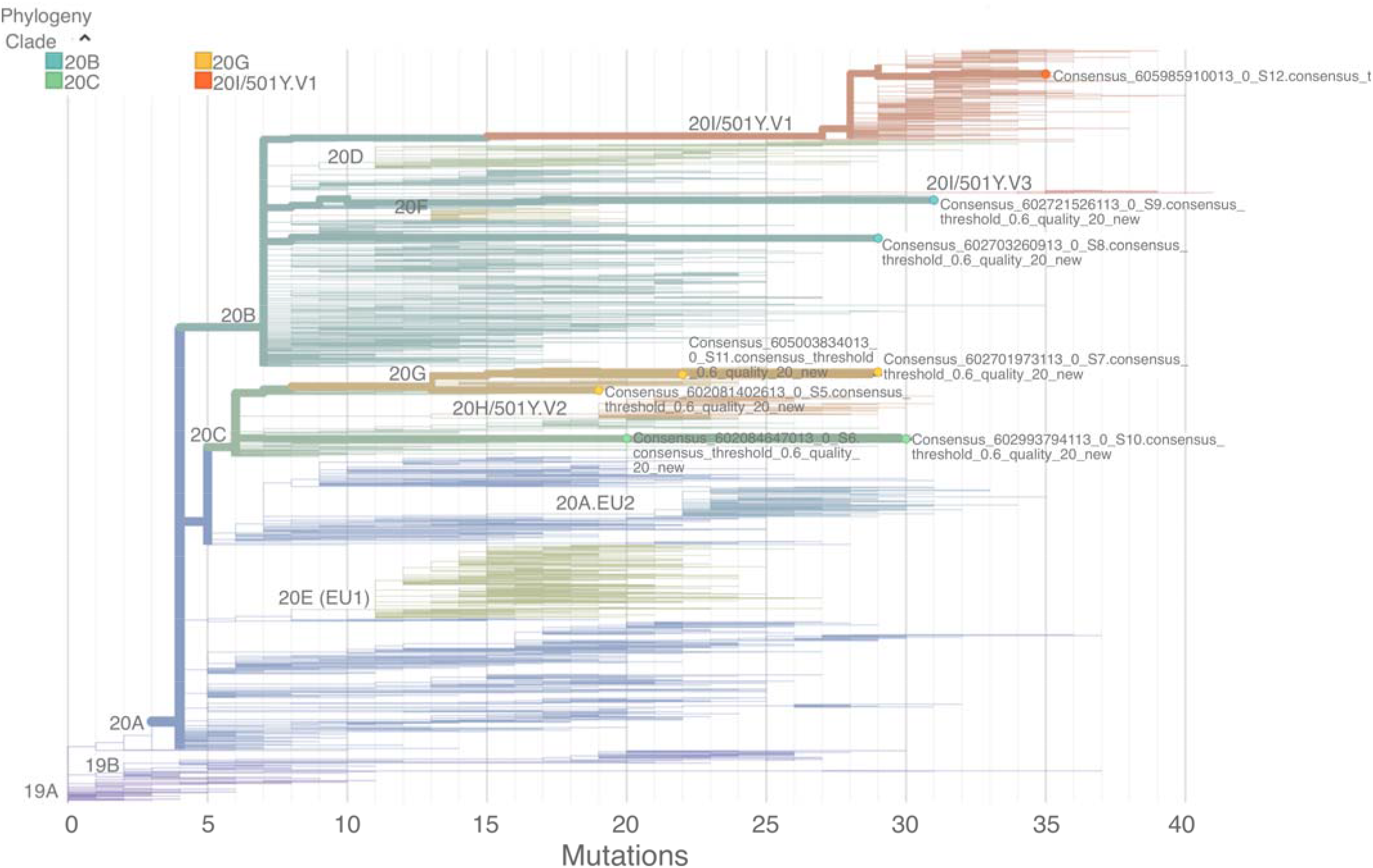
Phylogenetic analysis of the SARS-CoV-2 variants using Nextstrain clades. Figure shows that the methodologies described above are sensitive enough to resolve, not only large-scale genomic differences across different clades, but also differences within clades.

Phylogenetic relatedness may further contribute to the understanding of the pathogenic dynamics of SARS-CoV-2 and associated strains over time and link these genetic variations to specific geographical regions, which in turn can target genomic surveillance efforts.

## CONCLUSIONS

The present study evaluated tailed amplicon sequencing as a suitable approach for the detection of SARS-CoV-2 variants in the saliva of infected individuals collected using OMNIgene^®^•ORAL OME-505. Near complete genome sequences were obtained using the evaluated method, which in turn facilitated phylogenetic analysis with various SARS-CoV-2 variants. Near complete or complete genome sequences from SARS-CoV-2 variants continue to be invaluable in disease control and prevention efforts during the COVID-19 pandemic; thus, the evaluation of time-and cost-effective methods such as tailed amplicon sequencing is essential.

## Data Availability

SARS-CoV-2 sequence genomes are available on the Global Initiative on Sharing Avian Influenza Data (GISAID) site under IDs EPI_ISL_2156824, EPI_ISL_2156826, EPI_ISL_2156827, EPI_ISL_2156821, EPI_ISL_2156810, EPI_ISL_2156811, EPI_ISL_2156823, EPI_ISL_2156808.

## Author contributions

AG-data analysis, figure generation and manuscript draft editing. TSR-writing and editing of original manuscript draft. HLF-sample collection and screening for positive samples RI-conceptualization, and manuscript draft editing.

## Acknowledgements

We thank Mike Tayeb, Geoffrey M. Graham and Austin Udocor for reviewing the manuscript draft.

## Funding

No funding was received for this study.

## Informed Consent and Release Statement

Written informed consent and release statement has been obtained from the patient(s) to publish this paper.

## Institutional Review Board Statement

The study is exempt under 45 CFR § 46.104(d)(4), because the research involves the use of identifiable private information/biospecimens; and information, which may include information about biospecimens, is recorded by the investigator in such a manner that the identity of the human subjects cannot readily by ascertained directly or through identifiers linked to the subjects, the investigator does not contact the subjects, and the investigator will not re-identify subjects.

## Conflict of interest

AG and TSR are current employees of Diversigen Inc. RI is a current employee of DNA Genotek.

## Notes

### Author Declarations

Informed Consent and Release Statement Written informed consent and release statement has been obtained from the patient(s) to publish this paper. Institutional Review Board Statement The study is exempt under 45 CFR 46.104(d)(4), because the research involves the use of identifiable private information/biospecimens; and information, which may include information about biospecimens, is recorded by the investigator in such a manner that the identity of the human subjects cannot readily by ascertained directly or through identifiers linked to the subjects, the investigator does not contact the subjects, and the investigator will not re-identify subjects.

